# Anaemia in pregnancy in Bangladesh by the WHO 2024 haemoglobin cut-offs: a national biomarker survey and comparison with 2011

**DOI:** 10.64898/2026.10.20.26366027

**Authors:** S. M. Ashraf Ul Hoque, Md Khairul Islam, Mohammad Khalid Hasan, Mohiuddin Sharif, Fahima Sharmin Hossain, S M Raysul Haque, Kamran ul Baset

**Author notes:** Corresponding author: Mohammad Khalid Hasan, MBBS, MPH. Department of Public Health, Independent University Bangladesh (IUB), Plot 16 Aftab Uddin Ahmed Rd, Dhaka 1229, Bashundhara Residential Area, Dhaka 1245, Bangladesh.

## Abstract

**Background:** Anaemia in pregnancy is linked to low birthweight, preterm birth and maternal death. Bangladesh has had no publicly documented national haemoglobin survey of pregnant women since 2011, and WHO revised its cut-offs in 2024. We estimated national prevalence under these cut-offs, described haemoglobin across gestation, and compared 2025 with 2011.

**Methods:** We analysed venous haemoglobin from 1,933 of 2,711 eligible pregnant women in the nationally representative Bangladesh Multiple Indicator Cluster Survey 2025. We reanalysed capillary haemoglobin from 352 pregnant women in the Bangladesh Demographic and Health Survey 2011. Anaemia was haemoglobin below 11.0 g/dL in the first and third trimesters and below 10.5 g/dL in the second. Survey-weighted modified Poisson models estimated prevalence ratios (PR).

**Results:** In 2025, 53.2% (95% CI 50.6 to 55.8) of pregnant women were anaemic and 22.3% (20.2 to 24.5) had moderate or severe anaemia. A single 11.0 g/dL cut-off gave 60.7%. Prevalence was 42.1% in the first trimester and 67.7% in the third. Mean haemoglobin was lowest in month 6 and only 0.23 g/dL higher in months 8 and 9 than in months 5 and 6. Prevalence was similar across wealth, education and division. It was higher in Hindu women (adjusted PR 1.27, 95% CI 1.12 to 1.45) and slightly higher in urban women (1.13, 1.02 to 1.26). Under identical rules, prevalence was 42.9% (37.1 to 48.8) in 2011 (adjusted PR for 2025, 1.33, 1.15 to 1.54; 1.29, 1.11 to 1.50 with haemoglobin rounded as in 2011). The difference was concentrated in urban women (2.36, 1.62 to 3.42) rather than rural women (1.14, 0.97 to 1.35); the interaction had p<0.001. An assay offset of 0.4 g/dL removed the national difference but not the urban one.

**Conclusions:** About half of pregnant women in Bangladesh are anaemic, two in three by the third trimester, across all social groups. We found no evidence that anaemia has declined since 2011, and urban women appear to have lost their earlier advantage. Antenatal programmes need universal reach into late pregnancy, and surveys need harmonised haemoglobin methods.

## Introduction

Anaemia affects more than one in three pregnant women worldwide. A pooled analysis estimated a global prevalence of 36% (95% uncertainty interval 34 to 39) in 2019, with South Asia among the most affected regions [1,2]. Low maternal haemoglobin is associated with low birthweight, preterm birth, stillbirth, perinatal death and postpartum haemorrhage [3,4]. Severe anaemia is associated with about twice the odds of maternal death [5]. Halving anaemia among women of reproductive age is a World Health Assembly target extended to 2030, and few countries are on course to meet it [1].

The definition of anaemia in pregnancy changed in 2024. The World Health Organization (WHO) kept the 11.0 g/dL cut-off for the first and third trimesters. It set 10.5 g/dL for the second, when plasma volume expands fastest [6]. Pooled data from eight countries, including Bangladesh, support a lower second-trimester threshold [7]. Most national estimates still use a single 11.0 g/dL cut-off. Few have shown how much the new rule changes a national figure, or how haemoglobin moves across pregnancy in a national sample.

Bangladesh has lacked a documented national household survey estimate for 14 years. The Bangladesh Demographic and Health Survey (BDHS) 2011 found anaemia in 49.6% of 352 tested pregnant women, using the single cut-off [8]. Later BDHS rounds in 2014, 2017 to 2018 and 2022 did not measure haemoglobin, and the National Micronutrient Survey 2011 to 2012 sampled only non-pregnant, non-lactating women [9]. Anaemia in Bangladesh is not driven by iron deficiency alone. Thalassaemia traits are common, and iron in groundwater raises iron status in many areas [9 to 12]. Without current data, antenatal programmes cannot judge whether the burden has fallen or where it now concentrates. The Multiple Indicator Cluster Survey (MICS) 2025 measured venous haemoglobin in all eligible pregnant women for the first time [13]. A national low birthweight survey in 2015 also reported maternal haemoglobin, but the timing of measurement and the cut-off it used are not publicly documented [14].

We aimed to provide the first national household survey estimate of anaemia in pregnancy in Bangladesh since 2011, and the first by the WHO 2024 cut-offs. We first estimated prevalence and severity, and how much the trimester-specific rule changed the estimate against the single cut-off. We then described haemoglobin across the months of pregnancy and examined which women were most affected. Finally, we reanalysed the 2011 survey with identical rules to compare the two periods and to find where any change occurred.

## Methods

### Data sources and design

We used two nationally representative household surveys. The Bangladesh Bureau of Statistics conducted MICS 2025 with support from UNICEF [13,15]. The BDHS 2011 was conducted by the National Institute of Population Research and Training with ICF International [8]. Both used stratified two-stage cluster samples of households. We followed the STROBE guideline for cross-sectional studies [16].

### Participants and blood collection

In MICS 2025, every currently pregnant woman aged 15 to 49 years identified in an interviewed household was eligible for blood collection. Trained phlebotomists collected venous blood from consenting women between February and June 2025. Designated reference laboratories measured haemoglobin [13]. The survey report and public data do not name the assay. Of 2,711 eligible women, 1,940 gave blood, 1,934 had a valid result, and 1,933 had a recorded gestational age (S1 Fig).

In the BDHS 2011, haemoglobin was measured in ever-married women in a one-third subsample of households, from July to December 2011 [8]. Trained staff measured capillary haemoglobin with a portable HemoCue analyser. Of 1,079 currently pregnant women, 382 lived in selected households and 357 had a result. Five women, all aged 14 years, had a sampling weight of zero, leaving 352 for weighted analysis (S1 Fig).

### Haemoglobin and anaemia

The primary outcome was any anaemia by the WHO 2024 cut-offs [6]. These are haemoglobin below 11.0 g/dL in the first and third trimesters and below 10.5 g/dL in the second. Mild anaemia was 10.0 to 10.9 g/dL in the first and third trimesters and 9.5 to 10.4 g/dL in the second. Moderate anaemia was 7.0 g/dL up to the mild band, and severe anaemia was below 7.0 g/dL. Secondary outcomes were anaemia by a single 11.0 g/dL cut-off and mean haemoglobin. We used unadjusted haemoglobin in both surveys. Bangladesh is low-lying, and MICS did not record smoking by the woman herself.

MICS recorded the woman’s report of weeks of pregnancy. Half the values were multiples of four, which suggests conversion from months. We defined trimesters as weeks 1 to 13, 14 to 27 and 28 or more. To place both surveys on one scale, we converted weeks to months of pregnancy (weeks divided by four, rounded, from 1 to 9). The BDHS recorded completed months, which we grouped as months 1 to 3, 4 to 6 and 7 to 9.

### Characteristics

We chose covariates in advance from the literature on anaemia in South Asia [17 to 19]. They were trimester, age, number of previous births, education, household wealth quintile, urban or rural residence, division, religion and drinking water source. Water source (piped, tube well or other) served as a rough marker of groundwater iron exposure [10,11].

For the comparison across surveys, we harmonised education as 0 to 5, 6 to 10 and 11 or more years of schooling. We used the seven divisions of 2011 and merged Mymensingh, created in 2015, into Dhaka. Wealth quintiles are relative within each survey.

### Statistical analysis

Our analyses followed the three aims. All estimates accounted for strata, clusters and weights. In MICS 2025 we used the blood weight, which corrects for differences in testing between strata. We first estimated the prevalence of anaemia by severity, with 95% CIs from the logit transformation, and compared the two cut-off rules. We tested differences across groups with design-based chi-square tests.

For the second aim, we estimated mean haemoglobin and prevalence by month of pregnancy. Each woman was measured once, so these estimates compare women at different stages of pregnancy, not change within women. We fitted a restricted cubic spline of haemoglobin on weeks of pregnancy, with knots at 8, 16, 24, 32 and 38 weeks. We compared mean haemoglobin in months 8 and 9 with months 5 and 6. To describe which women were most affected, we estimated prevalence ratios (PR) with modified Poisson regression and a log link [20]. Each characteristic had its own model, adjusted for a minimal set of variables chosen in advance as plausible prior causes of that characteristic (S3 Table). Trimester was in every set because the cut-off depends on it. We also fitted one model with all characteristics, and linear models for mean haemoglobin (S3 Table). Moderate or severe anaemia was a secondary outcome in the same models, and we tested a linear trend across wealth quintiles. One woman lacked education data, so models used 1,932 women with complete data. These associations are descriptive and were not corrected for multiple comparisons.

For the third aim, we pooled the two surveys and normalised each survey’s weights to its sample size. We estimated crude PRs for 2025 against 2011, then PRs adjusted for trimester alone and for all harmonised covariates. Trimester adjustment removes differences in the trimester mix, which changes prevalence through the cut-off. We report the fully adjusted PR as the main comparison because age, previous births, education and residence also changed between surveys. We tested whether the difference varied by trimester, residence, education and wealth with interaction terms, and estimated stratum-specific PRs with subpopulation variance. We also compared adjusted mean haemoglobin by residence.

We ran several sensitivity analyses. We rounded MICS haemoglobin to one decimal, as recorded in the BDHS, for the 2025 estimate and for the comparison with 2011. We excluded weeks 12 to 15 and 26 to 29, where trimester is most uncertain, and separately excluded heaped weeks. We excluded haemoglobin below 6 or above 15 g/dL. We replaced the blood weight with the design weight, and with inverse probability of response weights. The response model included age, previous births, education, wealth, residence, division, trimester, religion and water source. Because assays differed, we added fixed offsets to 2025 values, from 0.10 to 0.45 g/dL in steps of 0.01 g/dL. We recorded the offset at which the 95% CI of the PR first included 1, and the offset at which the PR reached 1. We added the same offsets to the comparison by residence after seeing the residence interaction, so that analysis is post hoc. Analyses used Stata 17, and figures used R 4.6.1.

### Ethics

This study used publicly available data from the Bangladesh Multiple Indicator Cluster Survey (MICS) 2025 and the Bangladesh Demographic and Health Survey (BDHS) 2011. As a secondary data analysis, it did not require ethical clearance. The icddr,b Ethical Review Committee approved the MICS 2025 protocol, including blood specimen collection, in November 2024 (Protocol Reference: PR-24113). Mothers or legal guardians gave written informed consent before blood collection. Participants with severe anaemia or dangerously high heavy metal concentrations received immediate counselling and were referred through the Ministry of Health and Family Welfare for clinical management. The ICF International Institutional Review Board approved the BDHS 2011 procedures, and the National Ethics Review Committee of the Ministry of Health and Family Welfare also approved the protocol (NIPORT, Mitra and Associates, and ICF International, 2013). We accessed anonymized microdata for research purposes in line with the data sharing policies of BBS, UNICEF, and the DHS Program.

## Results

The 1,933 women analysed in 2025 had a mean age of 24.3 years and a mean gestation of 20.0 weeks (Table 1). Testing reached 71.2% (95% CI 69.2 to 73.2) of eligible women. It was lower among the richest women (55.7%), women with higher secondary education (60.5%), women in Dhaka division (62.2%) and women in the third trimester (67.1%) (S1 Table). Our estimates reproduced the published MICS 2025 and BDHS 2011 tables (S2 Table).

**Table 1.**
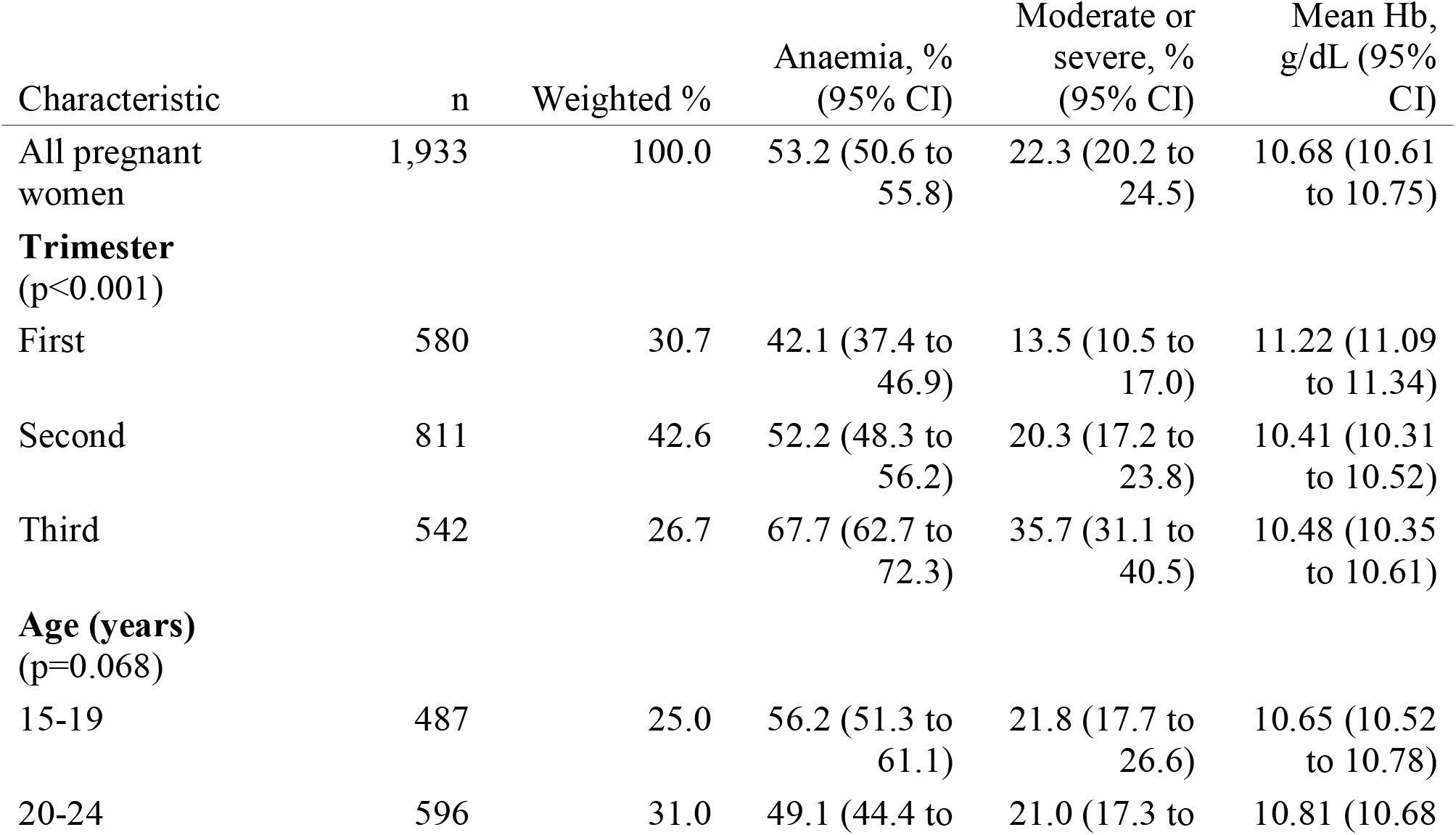
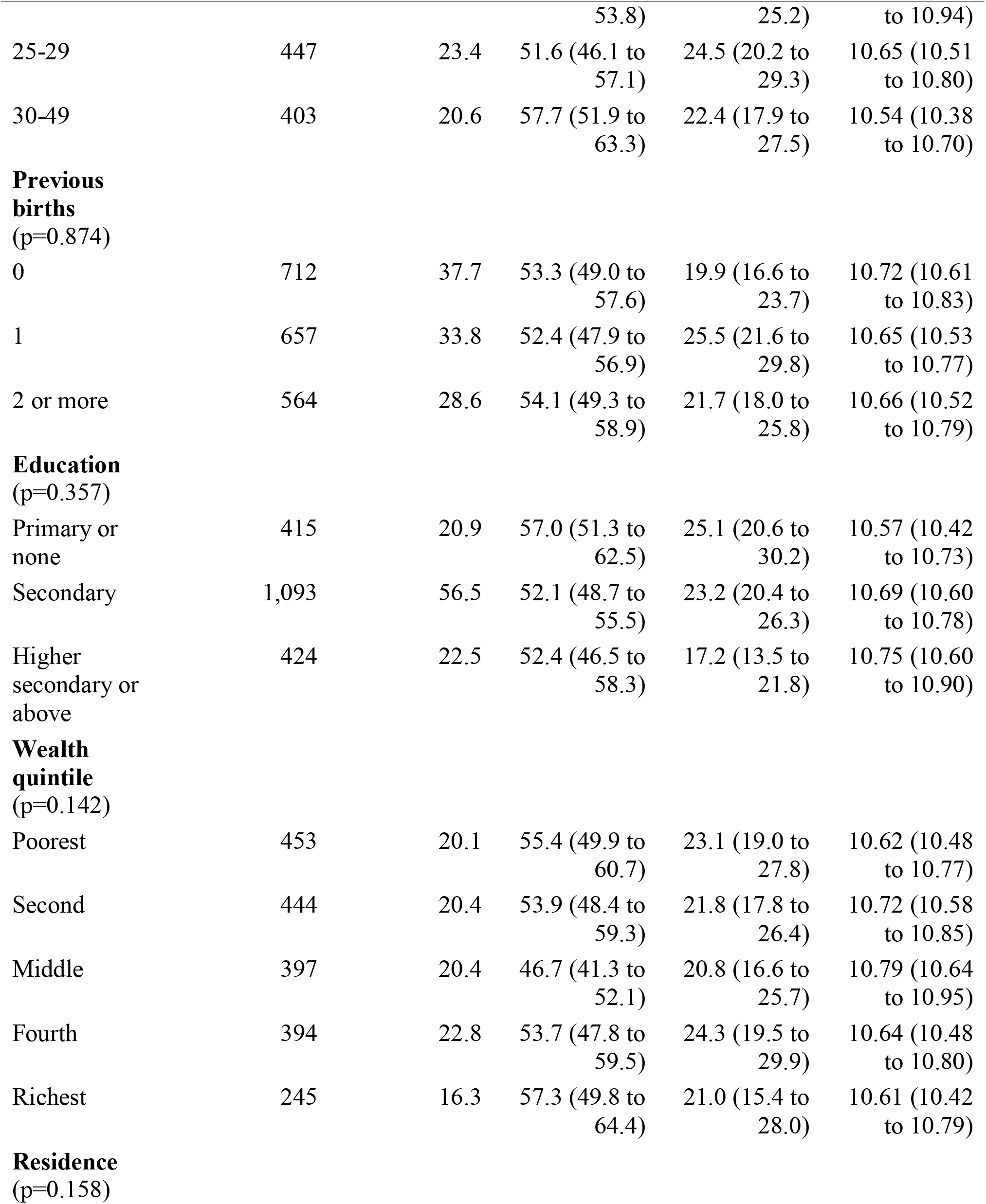
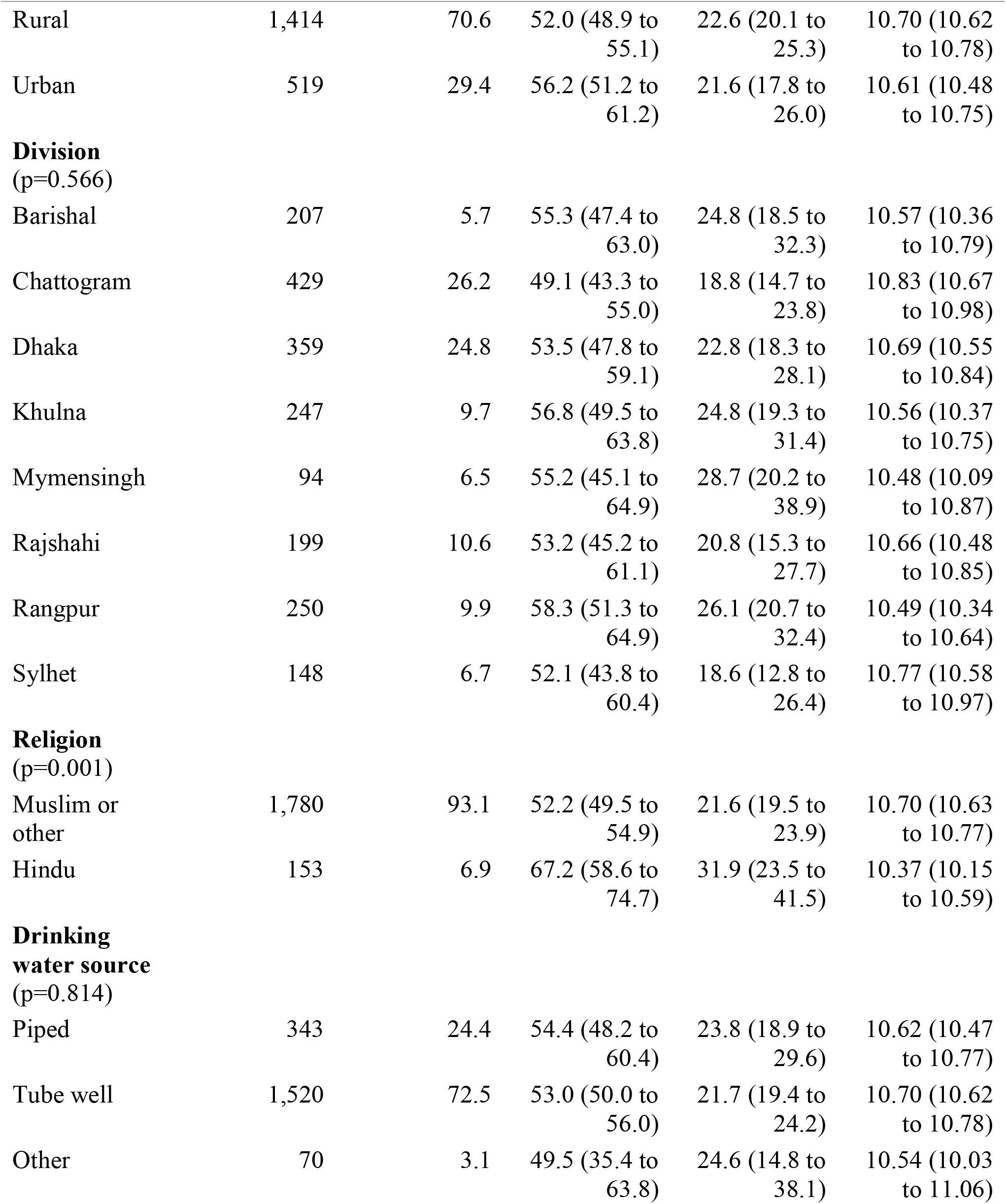
Characteristics of pregnant women and prevalence of anaemia by the WHO 2024 cut-offs, Bangladesh MICS 2025 (n=1,933). Percentages and means are survey-weighted with the blood weight; n is unweighted. p values are design-based chi-square tests of anaemia across categories. Moderate or severe: below 10.0 g/dL in the first and third trimesters and below 9.5 g/dL in the second. Education is missing for one woman.

In 2025, 53.2% (95% CI 50.6 to 55.8) of pregnant women had anaemia (Table 1). Mild anaemia affected 30.9% (28.6 to 33.4), moderate anaemia 21.6% (19.5 to 23.8) and severe anaemia 0.7% (0.4 to 1.4). Mean haemoglobin was 10.68 g/dL (10.61 to 10.75). A single 11.0 g/dL cut-off gave a prevalence of 60.7% (58.1 to 63.3). The 7.5 percentage point gap came entirely from second-trimester women with haemoglobin of 10.5 to 10.9 g/dL. In that trimester, prevalence was 52.2% by the WHO 2024 rule and 69.8% by the single cut-off.

Mean haemoglobin was 11.22 g/dL (11.09 to 11.34) in the first trimester and lowest in month 6, at 10.25 g/dL (10.04 to 10.46) (Figure 1A). Women in months 8 and 9 had a mean 0.23 g/dL (0.02 to 0.45) higher than women in months 5 and 6. Mean haemoglobin was similar in the second and third trimesters (10.41 and 10.48 g/dL). The cut-off returns to 11.0 g/dL in the third trimester. Prevalence was therefore 42.1% in the first trimester, 52.2% in the second and 67.7% in the third (Figure 1B). Moderate or severe anaemia was 13.5% in the first trimester and 35.7% in the third.

**Figure 1.**
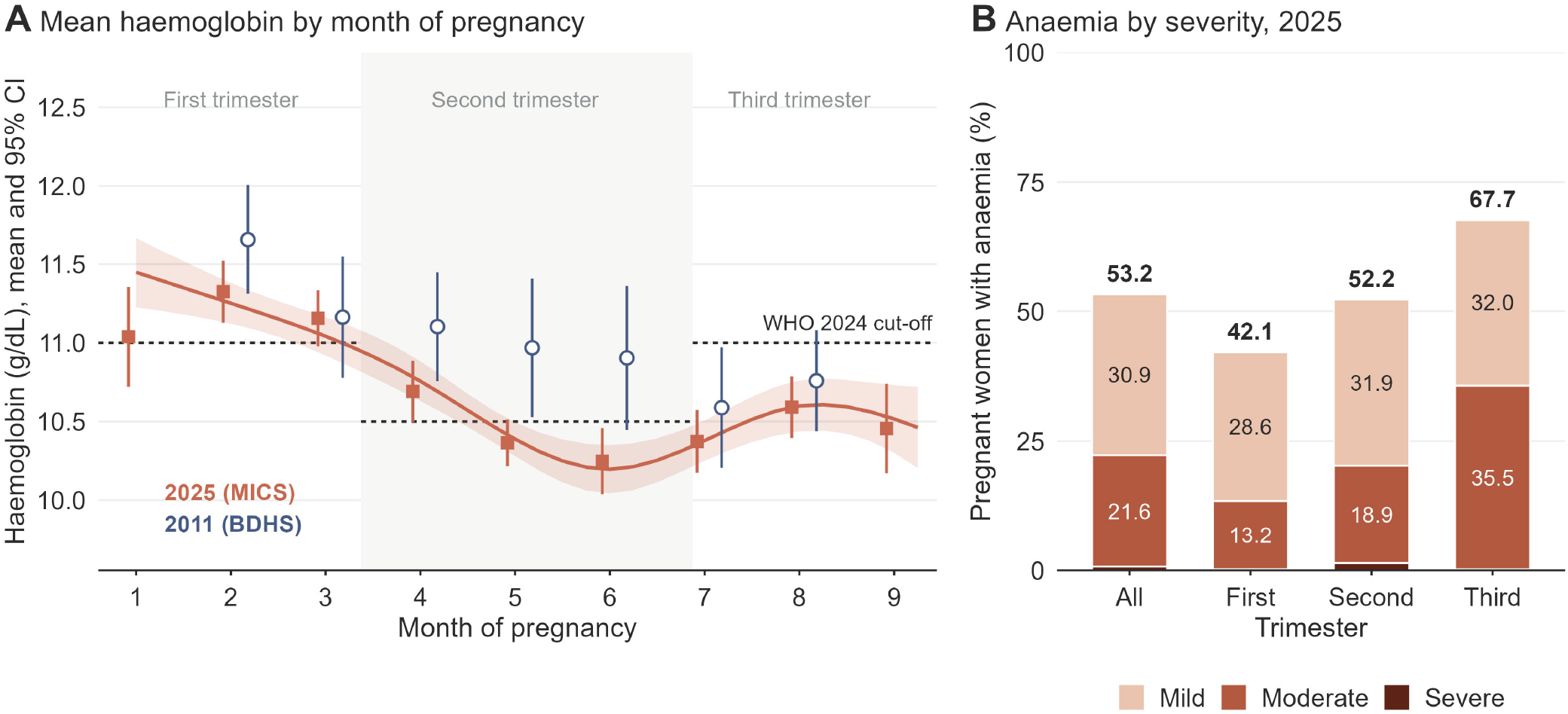
Haemoglobin across pregnancy and anaemia by severity. (A) Mean haemoglobin (95% CI) by month of pregnancy in MICS 2025 (squares) and BDHS 2011 (circles), with a restricted cubic spline for 2025 (line and band). MICS weeks of pregnancy were divided by four. Each month contains different women, so the curve compares stages of pregnancy, not change within women. BDHS months with fewer than 25 women (months 1 and 9) are not shown. Dashed lines mark the WHO 2024 cut-offs. (B) Prevalence of mild, moderate and severe anaemia by trimester in 2025; totals above bars. Severe anaemia (0.7% overall) is the thin dark band.

Prevalence was similar across wealth quintiles, education and divisions (Table 1; Figure 2). Divisional estimates ranged from 49.1% in Chattogram to 58.3% in Rangpur (p=0.566), and quintile estimates from 46.7% to 57.3% (p=0.142). Compared with the first trimester, prevalence was higher in the second (PR 1.24, 95% CI 1.08 to 1.42) and third (1.61, 1.41 to 1.84) (Figure 3). Hindu women had higher prevalence than Muslim and other women (67.2% against 52.2%; adjusted PR 1.27, 1.12 to 1.45). Urban women had slightly higher prevalence than rural women (56.2% against 52.0%; adjusted PR 1.13, 1.02 to 1.26). The crude ratio was 1.08 (0.97 to 1.21).

**Figure 2.**
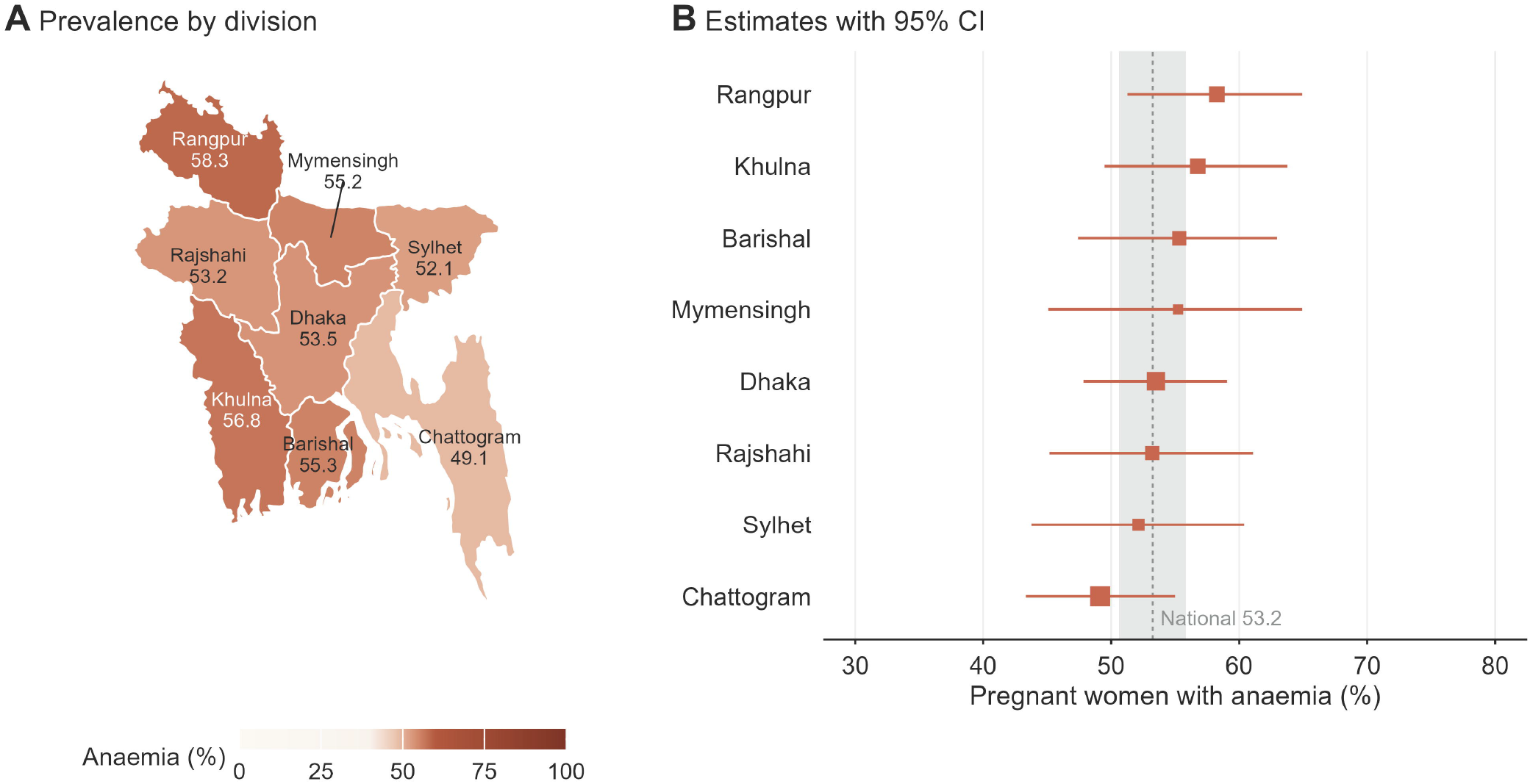
Anaemia in pregnancy by division, MICS 2025. (A) Prevalence by division on a 0 to 100% scale, with colour steps concentrated over the observed range. (B) Divisional estimates with 95% CI; the dashed line and grey band show the national estimate and its 95% CI. Square size reflects the number of women. p=0.566 for differences between divisions.

**Figure 3.**
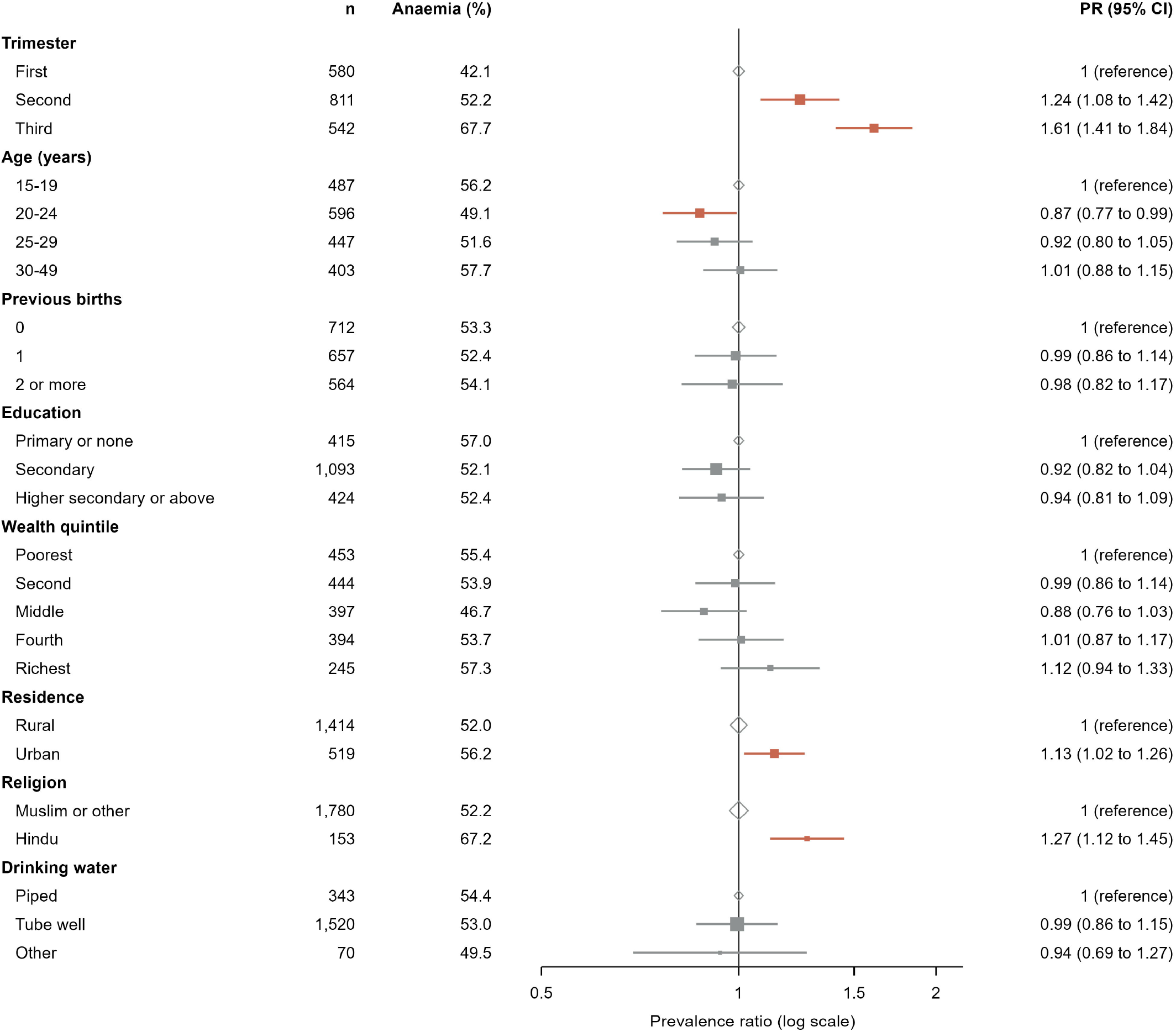
Characteristics associated with anaemia in pregnancy, MICS 2025. Prevalence ratios (95% CI) from survey-weighted modified Poisson models (n=1,932). Each characteristic comes from its own model, adjusted for a minimal set of prior variables listed in S3 Table; trimester is unadjusted. Ratios are descriptive and not mutually adjusted. Square size reflects the number of women. Terracotta marks intervals that exclude 1.

Wealth showed no linear trend (p=0.318). Women aged 20 to 24 years had lower prevalence than those aged 15 to 19 (adjusted PR 0.87, 0.77 to 0.99). The overall test for age had p=0.100.

Previous births, education, division and water source showed no clear association (S3 Table). For moderate or severe anaemia, a secondary outcome, prevalence was lower with higher secondary education or above (adjusted PR 0.67, 0.49 to 0.91). It was higher in Hindu women (1.47, 1.08 to 1.99). Estimates from one mutually adjusted model were similar (S3 Table).

Under identical rules, prevalence was 42.9% (95% CI 37.1 to 48.8) in 2011 and 53.2% (50.6 to 55.9) in 2025 (Table 2; Figure 4). The PR for 2025 against 2011 was 1.24 (1.07 to 1.44) crude, 1.28 (1.11 to 1.47) adjusted for trimester and 1.33 (1.15 to 1.54) fully adjusted. With 2025 haemoglobin rounded to one decimal, as in 2011, prevalence in 2025 was 51.0% and the fully adjusted PR was 1.29 (1.11 to 1.50). Moderate or severe anaemia rose from 15.1% to 22.3% (adjusted PR 1.77, 1.33 to 2.36). Mean haemoglobin was 0.42 g/dL (0.25 to 0.59) lower in 2025 after adjustment. Women in 2025 were more educated, slightly older and more often urban than in 2011 (S5 Table).

**Table 2.**
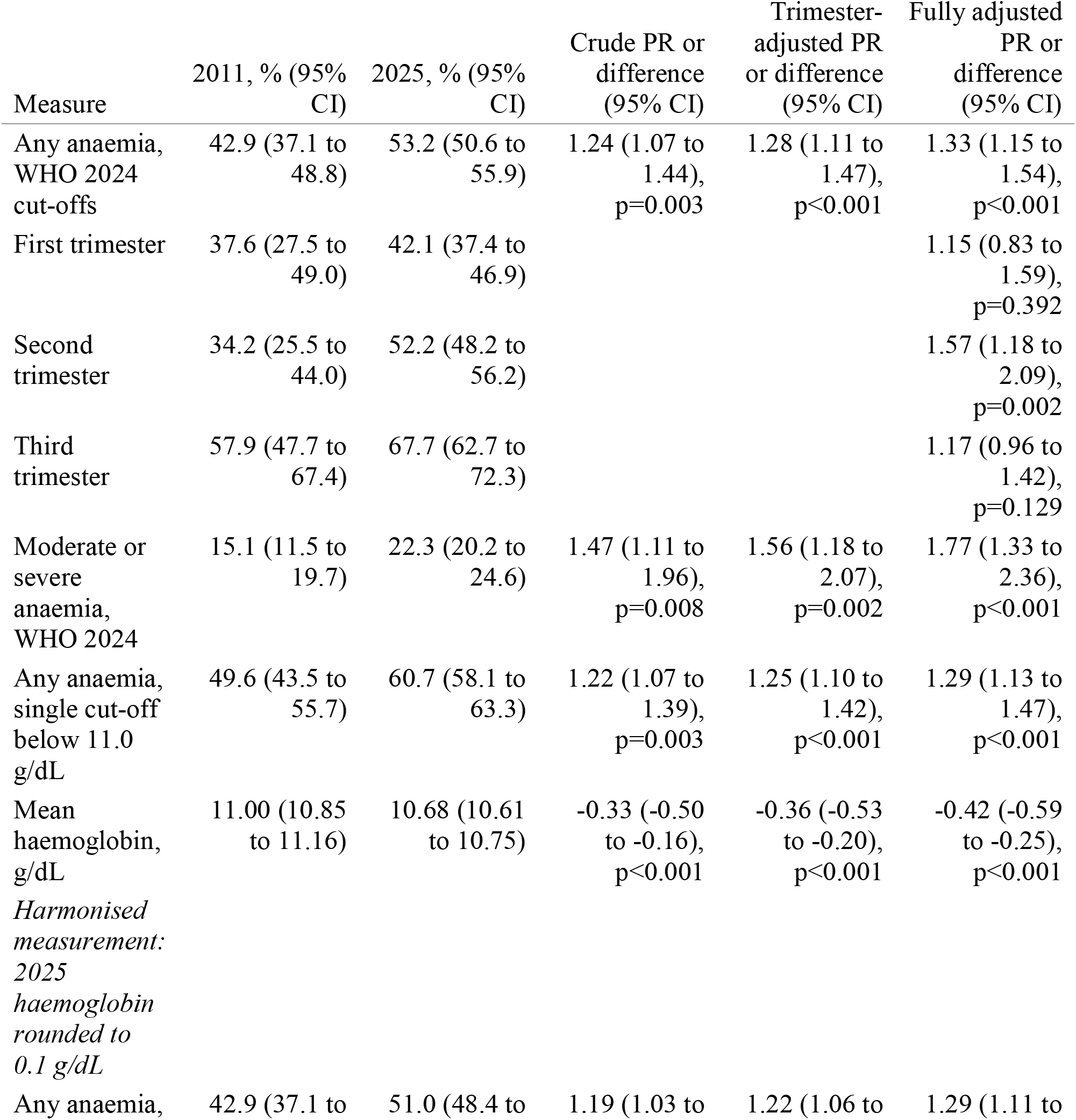
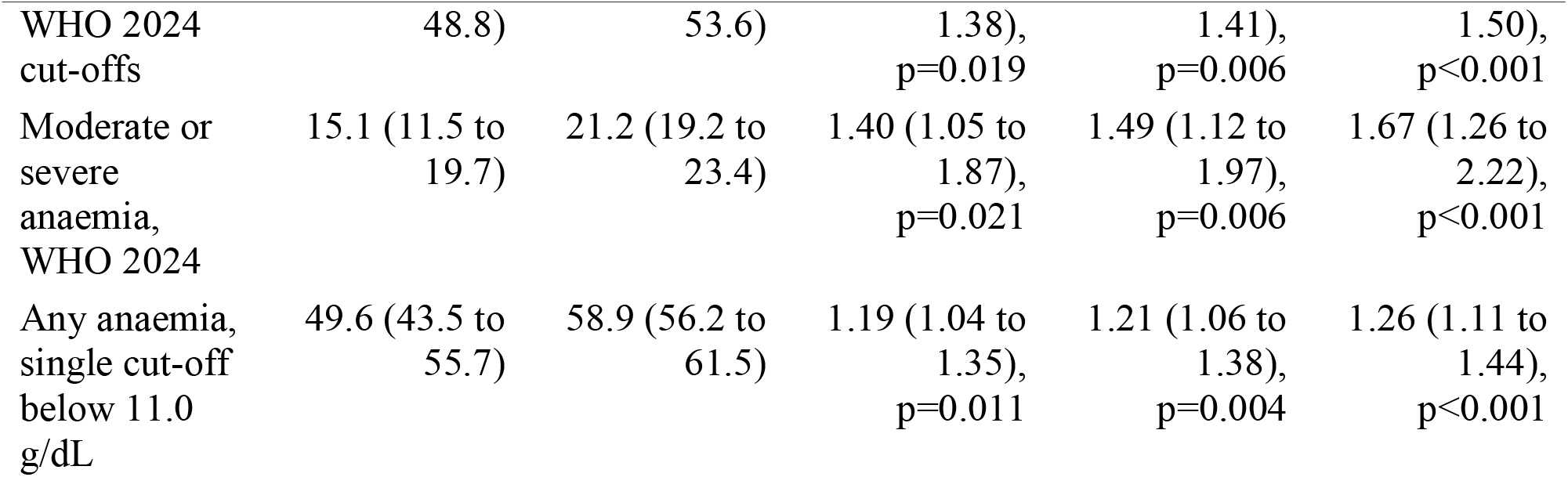
Anaemia in pregnant women in 2011 and 2025 under identical definitions. BDHS 2011 (n=352, capillary haemoglobin) and MICS 2025 (n=1,933, venous haemoglobin). Estimates use each survey’s weights within a pooled design, so 2025 intervals may differ from Table 1 in the last digit. PR, prevalence ratio for 2025 against 2011; for mean haemoglobin, the difference in g/dL. Fully adjusted models include trimester, age, previous births, education in years, residence, division (2011 boundaries) and relative wealth quintile; trimester-specific PRs omit the trimester term and use subpopulation variance. Interaction between survey and trimester p=0.231. Harmonised-measurement rows round 2025 haemoglobin to 0.1 g/dL, as recorded in 2011; the rounded fully adjusted mean difference was −0.42 g/dL (−0.59 to −0.25).

**Figure 4.**
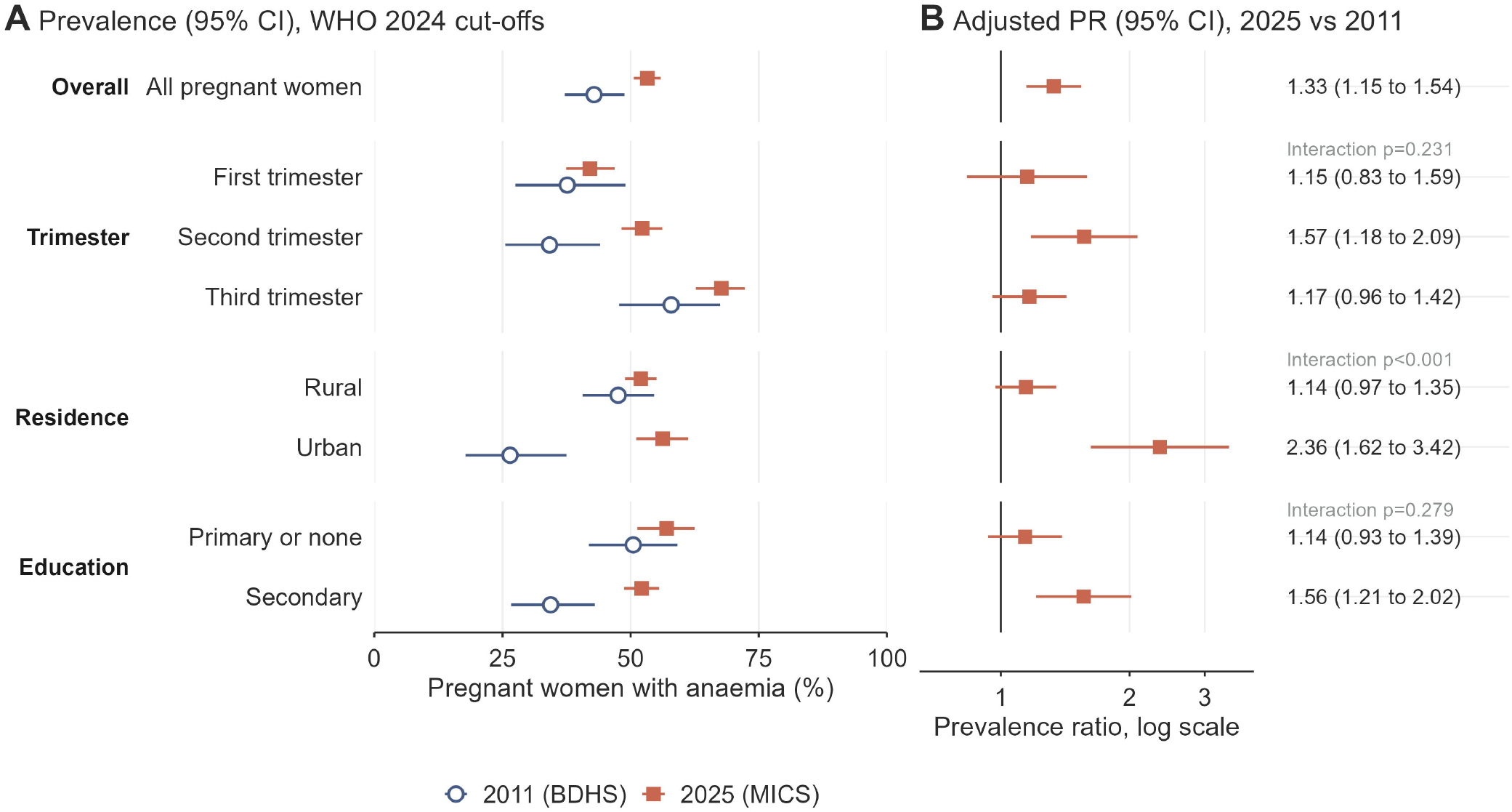
Anaemia in pregnant women in 2011 and 2025 under identical WHO 2024 cut-offs. (A) Prevalence with 95% CI by survey. (B) Adjusted prevalence ratio for 2025 against 2011, with p values for interaction between survey and each characteristic. Stratum-specific ratios come from subpopulation models. Education above secondary is omitted because only 20 women in 2011 had 11 or more years of schooling.

The difference sat mainly in urban women. Urban prevalence was 26.5% (17.8 to 37.5) among 107 women in 2011 and 56.2% (51.1 to 61.2) in 2025 (adjusted PR 2.36, 1.62 to 3.42). Rural prevalence was 47.6% (40.7 to 54.6) and 52.0% (48.9 to 55.1) (adjusted PR 1.14, 0.97 to 1.35). The interaction between survey and residence had p<0.001. In 2011, urban women had less anaemia than rural women (adjusted PR 0.65, 0.43 to 0.97); in 2025 they did not (1.09, 0.98 to 1.22). Adjusted mean haemoglobin was 0.91 g/dL (0.60 to 1.22) lower in 2025 than in 2011 in urban women. In rural women it was 0.27 g/dL (0.07 to 0.47) lower (interaction p<0.001). The PR also appeared larger in women with secondary education (1.56 against 1.14; interaction p=0.279) and in richer households (1.50 against 1.15; interaction p=0.064) (S4 Table).

National prevalence ranged from 51.0% to 53.9% across sensitivity analyses (S4 Table). The response-weighted estimate was 53.9% (51.2 to 56.5). Rounding MICS haemoglobin to one decimal gave 51.0%. Adding an offset to 2025 values reduced the adjusted PR against 2011. Its 95% CI first included 1 at an offset of 0.19 g/dL (PR 1.16, 1.00 to 1.36), and the PR reached 1.0 at 0.38 g/dL (1.00, 0.85 to 1.17). With a 0.4 g/dL offset, the PR was 1.66 (1.12 to 2.46) in urban women and 0.86 (0.72 to 1.03) in rural women (interaction p=0.002). Associations in 2025 changed little with response weights or without weights (S3 Table).

## Discussion

About half of pregnant women in Bangladesh were anaemic in 2025 by the WHO 2024 cut-offs, and two in three by the third trimester. Mean haemoglobin was lowest in the sixth month and only slightly higher later in pregnancy. Anaemia was spread almost evenly across wealth, education and divisions, with slightly more in urban women. Compared with 2011 under identical rules, prevalence was higher, and the urban advantage seen in 2011 had disappeared.

Our national figure updates the only earlier estimate, from the BDHS 2011. By the single 11.0 g/dL cut-off used in 2011, prevalence was 60.7% in 2025 against 49.6% in 2011 [8]. The 2025 figure also exceeds the global estimate of 36% for 2019 [1]. By the single cut-off, neighbouring countries report lower or similar figures [21]. These are 52.2% in India in 2019 to 2021, 32.7% in Nepal in 2022 and 56.9% in Myanmar in 2015 to 2016. These surveys used capillary blood and adjusted for altitude and smoking, so comparisons are approximate.

The WHO 2024 rule lowered the national estimate by 7.5 points, all in the second trimester. Pooled data from healthy women support this change [7]. Programme reports that switch rules without saying so will show a spurious fall. Both figures should therefore be reported during the transition.

The pattern across gestation is the clearest finding. Haemoglobin normally falls in the second trimester as plasma volume expands, then partly recovers [6]. Our data are cross-sectional and compare women at different stages. Women in months 8 and 9 had only 0.23 g/dL more haemoglobin than women in months 5 and 6. The WHO cut-off rises by 0.5 g/dL between these trimesters, which assumes more recovery than we observed [6]. Most women in the third trimester were therefore below 11.0 g/dL. Moderate or severe anaemia reached 35.7% in the third trimester, close to delivery, when blood loss is most dangerous [3,5]. The small difference is consistent with unmet iron or other nutrient needs late in pregnancy, but MICS did not measure iron status. In the BDHS 2017 to 2018, only 21.7% of women with a recent birth had taken iron and folic acid for the full 180 days [22].

The flat gradient by wealth, education and division points to factors that cut across social position and place. Iron deficiency was uncommon among non-pregnant Bangladeshi women in national data, and groundwater iron is linked to higher iron status [9 to 11]. An estimated 10.9% to 13.3% of people carry thalassaemia or haemoglobin E traits [12], which lower haemoglobin regardless of income. Pregnancy adds iron demands that diet and groundwater may not meet even in richer homes. The higher prevalence in Hindu women matches findings among non-pregnant women in the BDHS 2011 [17,18]. Diets with less animal-source food are a plausible but untested explanation. Moderate or severe anaemia was less common among the most educated women, so social position may matter more for severity than for any anaemia.

We cannot confirm a national rise with certainty, but we found no sign of a fall. The two surveys used different blood sources, devices and seasons. Method comparison studies disagree on direction. Capillary HemoCue readings were lower than venous analyser readings in India, but HemoCue gave less anaemia than an analyser in another Indian study [23,24]. Paired surveys found no consistent venous and capillary difference among non-pregnant women [25]. An offset of about 0.2 g/dL would make the national difference compatible with no change, and about 0.4 g/dL would remove it. It is therefore the size of measurement error, not its existence, that matters for the national trend. Only an offset above about 0.4 g/dL would point towards a fall.

The urban finding does not depend on a uniform assay offset. An offset applies to urban and rural women alike, yet the urban difference survived a 0.4 g/dL offset. It has two other weaknesses. The 2011 urban estimate rests on 107 women, and the 2011 urban advantage had p=0.037. Urban boundaries also changed between the surveys as towns grew and new municipalities formed. Newly urban areas may have brought poorer, more rural households into the urban group, which alone could erase the earlier advantage. In 2011, urban pregnant women had about a third less anaemia than rural women; in 2025 they had the same. Our data cannot explain this change. Changing urban diets, crowding and migration of poorer women to cities are possible drivers that need testing. Water source was not associated with anaemia in 2025, so a shift from tube wells to piped water seems unlikely to explain it.

Because anaemia was common in every group, targeting by wealth or region would miss most affected women. Antenatal care should offer iron-containing supplements to all pregnant women and support adherence through the third trimester [26]. In a Bangladeshi trial, multiple micronutrient supplements reduced preterm birth (RR 0.85) and low birthweight (RR 0.88) compared with iron and folic acid [27]. WHO recommends them in the context of rigorous research [28]. Haemoglobin testing in the third trimester could identify women needing treatment before delivery. Urban antenatal services deserve particular attention. Future national surveys should use venous blood, document the assay and include iron, inflammation and haemoglobin disorder markers.

To our knowledge, this is the first national household survey estimate of anaemia in pregnancy in Bangladesh since 2011, and the first by the WHO 2024 cut-offs. The 2015 national low birthweight survey reported maternal haemoglobin, but its timing and cut-off are not documented, so we could not compare with it [14]. Venous sampling in a national probability sample reduces measurement error compared with capillary drops. We reproduced the published tables and applied identical rules to the 2011 data.

Several limitations apply. First, 29% of eligible women gave no valid sample, and the richest women, the most educated and women in Dhaka division were tested less. The blood weight corrects for differences between strata, and response weights within strata changed prevalence by less than one point. The higher response-weighted estimate suggests that, if anything, we slightly underestimated prevalence. Bias would remain if testing depended on haemoglobin itself, in an unknown direction. Second, gestational age was self-reported and often rounded to months, so trimester may be misclassified near the boundaries. Excluding those weeks lowered prevalence by 1.3 points, so misclassification may slightly inflate the estimate. Third, the comparison with 2011 rests on different assays, seasons and urban boundaries, and on a small 2011 sample with wide intervals. The direction of assay bias is unknown; our offset analyses show how large it would need to be to explain the national difference. Fourth, we lacked data on iron status, inflammation, haemoglobin disorders, diet and supplement use in the current pregnancy. Fifth, the design is cross-sectional, so associations do not show causes. The Hindu subgroup was small, and some associations may be chance findings among many comparisons.

## Conclusions

About half of pregnant women in Bangladesh were anaemic in 2025 by the WHO 2024 cut-offs. Prevalence reached two in three by the third trimester and was similar across social groups and regions. We found no evidence that anaemia had declined since 2011, and urban women appeared to have lost their earlier advantage. Antenatal programmes need universal coverage that continues into late pregnancy, and national surveys need harmonised haemoglobin methods to track progress.

## Declarations

### Data availability

MICS 2025 data are available on request from UNICEF MICS (https://mics.unicef.org). BDHS 2011 data are available on request from the DHS Program (https://dhsprogram.com).

### Author contributions

SMAUH conceived and designed the study, developed the methodology, curated the data, carried out the formal analysis, prepared the software and figures, and wrote the original draft. KUB supervised the study and critically reviewed the manuscript. MKH developed the methodology, wrote the original draft, administered the project and the journal submission. MKI, MS, FSH and SMRH critically reviewed the results and their interpretation. All authors read and approved the final manuscript for submission.

## Acknowledgements

We thank the Bangladesh Bureau of Statistics and UNICEF for access to the data.

---

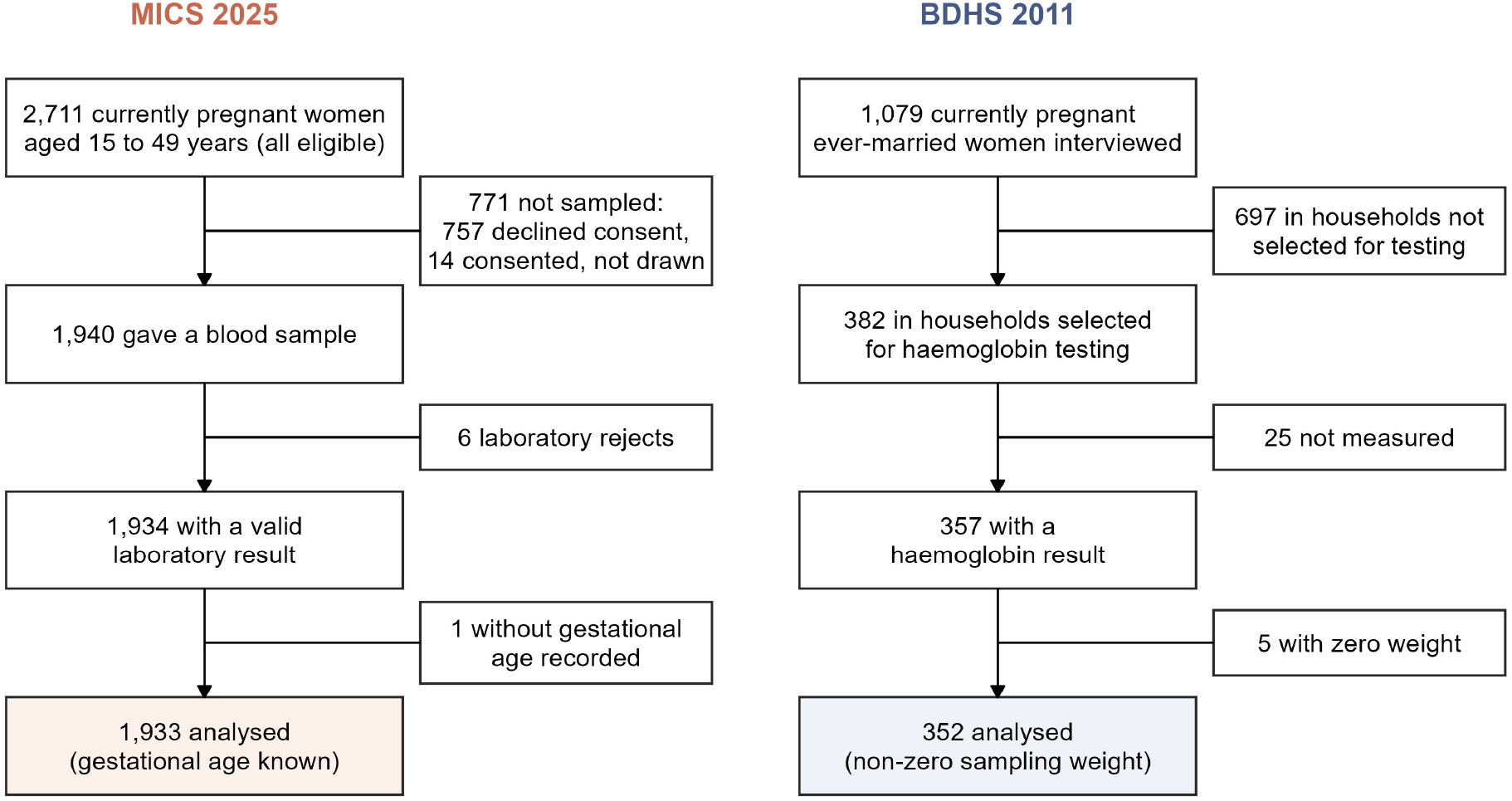

## References

1. Stevens GA, Paciorek CJ, Flores-Urrutia MC, et al. National, regional, and global estimates of anaemia by severity in women and children for 2000-19: a pooled analysis of population-representative data. Lancet Glob Health 2022;10(5):e627–39. doi:10.1016/S2214-109X(22)00084-5

2. GBD 2021 Anaemia Collaborators. Prevalence, years lived with disability, and trends in anaemia burden by severity and cause, 1990–2021: findings from the Global Burden of Disease Study 2021. Lancet Haematol 2023;10(9):e713–34. doi:10.1016/S2352-3026(23)00160-6

3. Young MF, Oaks BM, Rogers HP, et al. Maternal low and high hemoglobin concentrations and associations with adverse maternal and infant health outcomes: an updated global systematic review and meta-analysis. BMC Pregnancy Childbirth 2023;23(1):264. doi:10.1186/s12884-023-05489-6

4. Rahman MM, Abe SK, Rahman MS, et al. Maternal anemia and risk of adverse birth and health outcomes in low- and middle-income countries: systematic review and meta-analysis. Am J Clin Nutr 2016;103(2):495–504. doi:10.3945/ajcn.115.107896

5. Daru J, Zamora J, Fernández-Félix BM, et al. Risk of maternal mortality in women with severe anaemia during pregnancy and post partum: a multilevel analysis. Lancet Glob Health 2018;6(5):e548–54. doi:10.1016/S2214-109X(18)30078-0

6. World Health Organization. Guideline on haemoglobin cutoffs to define anaemia in individuals and populations. Geneva: WHO; 2024.

7. Braat S, Fielding KL, Han J, et al. Haemoglobin thresholds to define anaemia from age 6 months to 65 years: estimates from international data sources. Lancet Haematol 2024;11(4):e253–64. doi:10.1016/S2352-3026(24)00030-9

8. National Institute of Population Research and Training (NIPORT), Mitra and Associates, ICF International. Bangladesh Demographic and Health Survey 2011. Dhaka and Calverton: NIPORT, Mitra and Associates, and ICF International; 2013.

9. Rahman S, Ahmed T, Rahman AS, et al. Determinants of iron status and Hb in the Bangladesh population: the role of groundwater iron. Public Health Nutr 2016;19(10):1862–74. doi:10.1017/S1368980015003651

10. Merrill RD, Shamim AA, Ali H, et al. Iron status of women is associated with the iron concentration of potable groundwater in rural Bangladesh. J Nutr 2011;141(5):944–9. doi:10.3945/jn.111.138628

11. Merrill RD, Shamim AA, Ali H, et al. High prevalence of anemia with lack of iron deficiency among women in rural Bangladesh: a role for thalassemia and iron in groundwater. Asia Pac J Clin Nutr 2012;21(3):416–24.

12. Hossain MS, Islam F, Akhter S, et al. Thalassemia in Bangladesh: progress, challenges, and a strategic blueprint for prevention. Orphanet J Rare Dis 2025;20(1):358. doi:10.1186/s13023-025-03744-x

13. Bangladesh Bureau of Statistics (BBS), UNICEF Bangladesh. Progotir Pathey, Bangladesh Multiple Indicator Cluster Survey 2025, Survey Findings Report. Dhaka: BBS and UNICEF Bangladesh; 2026.

14. National Nutrition Services, Institute of Public Health Nutrition. National Low Birth Weight Survey 2015, Bangladesh. Dhaka: IPHN; 2017.

15. Khan S, Hancioglu A. Multiple Indicator Cluster Surveys: delivering robust data on children and women across the globe. Stud Fam Plann 2019;50(3):279–86. doi:10.1111/sifp.12103

16. von Elm E, Altman DG, Egger M, et al. The Strengthening the Reporting of Observational Studies in Epidemiology (STROBE) statement: guidelines for reporting observational studies. Lancet 2007;370(9596):1453–7.

17. Kamruzzaman M, Rabbani MG, Saw A, et al. Differentials in the prevalence of anemia among non-pregnant, ever-married women in Bangladesh: multilevel logistic regression analysis of data from the 2011 Bangladesh Demographic and Health Survey. BMC Womens Health 2015;15:54. doi:10.1186/s12905-015-0211-4

18. Jana A, Chattopadhyay A, Saha UR. Identifying risk factors in explaining women’s anaemia in limited resource areas: evidence from West Bengal of India and Bangladesh. BMC Public Health 2022;22(1):1433. doi:10.1186/s12889-022-13806-5

19. Rahman MA, Rahman MS, Aziz Rahman M, et al. Prevalence of and factors associated with anaemia in women of reproductive age in Bangladesh, Maldives and Nepal: evidence from nationally-representative survey data. PLoS One 2021;16(1):e0245335. doi:10.1371/journal.pone.0245335

20. Zou G. A modified Poisson regression approach to prospective studies with binary data. Am J Epidemiol 2004;159(7):702–6. doi:10.1093/aje/kwh090

21. ICF. The DHS Program STATcompiler: anaemia in pregnant women (India 2019-21, Nepal 2022, Myanmar 2015-16). https://www.statcompiler.com (accessed 13 October 2026).

22. Sanin KI, Alam Shaun M, Rita RS, et al. What makes Bangladeshi pregnant women more compliant to iron-folic acid supplementation: a nationally representative cross-sectional survey result. Nutrients 2023;15(6):1512. doi:10.3390/nu15061512

23. Reddy VS, Reddy SN, Mahajan H, et al. Comparison of hemoglobin measurements from venous and capillary blood from the same individual using HemoCue 301 and automated hematology analyzer in a cross-sectional community-based study in India. Am J Clin Nutr 2026;123(1):101119. doi:10.1016/j.ajcnut.2025.11.009

24. Fothergill A, Crider KS, Johnson CB, et al. Comparison of anemia screening methods using paired venous samples in women of reproductive age in Southern India. J Nutr 2022;152(12):2978–92. doi:10.1093/jn/nxac218

25. Stevens GA, Flores-Urrutia MC, Rogers LM, et al. Associations between type of blood collection, analytical approach, mean haemoglobin and anaemia prevalence in population-based surveys: a systematic review and meta-analysis. J Glob Health 2022;12:04088. doi:10.7189/jogh.12.04088

26. World Health Organization. WHO recommendations on antenatal care for a positive pregnancy experience. Geneva: WHO; 2016.

27. West KP Jr, Shamim AA, Mehra S, et al. Effect of maternal multiple micronutrient vs iron-folic acid supplementation on infant mortality and adverse birth outcomes in rural Bangladesh: the JiVitA-3 randomized trial. JAMA 2014;312(24):2649–58. doi:10.1001/jama.2014.16819

28. World Health Organization. WHO antenatal care recommendations for a positive pregnancy experience. Nutritional interventions update: multiple micronutrient supplements during pregnancy. Geneva: WHO; 2020.

