## Supplementary material for "Anaemia in pregnancy in Bangladesh by the WHO 2024 haemoglobin cut-offs: a national biomarker survey and comparison with 2011": Supplement_Draft_v2.docx

S1 File. Supplementary tables and figure

### S1 Table. Blood testing among eligible pregnant women, MICS 2025

Percentage of the 2,711 eligible women with a valid haemoglobin result, weighted with the design weight. p values are design-based chi-square tests.

| Group | Characteristic | Eligible, n | Tested, % (95% CI) | p (design-based) |
| --- | --- | --- | --- | --- |
|  | All eligible women | 2,711 | 71.2 (69.2 to 73.2) |  |
| Age (years) | 15-19 | 686 | 71.7 (67.7 to 75.4) | p=0.497 |
|  | 20-24 | 860 | 69.5 (65.7 to 73.0) |  |
|  | 25-29 | 632 | 70.8 (66.4 to 74.8) |  |
|  | 30-49 | 533 | 74.0 (69.3 to 78.2) |  |
| Previous births | 0 | 1,071 | 67.1 (63.7 to 70.3) | p=0.004 |
|  | 1 | 898 | 73.6 (70.1 to 76.8) |  |
|  | 2 or more | 742 | 74.5 (70.5 to 78.1) |  |
| Education | Primary or none | 520 | 79.8 (75.2 to 83.7) | p<0.001 |
|  | Secondary | 1,494 | 73.5 (70.8 to 76.0) |  |
|  | Higher secondary or above | 696 | 60.5 (56.0 to 64.8) |  |
| Wealth quintile | Poorest | 551 | 83.2 (79.0 to 86.6) | p<0.001 |
|  | Second | 584 | 76.0 (71.7 to 79.7) |  |
|  | Middle | 563 | 72.2 (67.6 to 76.4) |  |
|  | Fourth | 564 | 70.0 (65.1 to 74.6) |  |
|  | Richest | 449 | 55.7 (50.4 to 60.8) |  |
| Residence | Rural | 1,955 | 72.3 (70.0 to 74.6) | p=0.101 |
|  | Urban | 756 | 68.5 (64.4 to 72.4) |  |
| Division | Barishal | 238 | 87.1 (81.4 to 91.2) | p<0.001 |
|  | Chattogram | 650 | 67.3 (63.0 to 71.4) |  |
|  | Dhaka | 587 | 62.2 (57.4 to 66.7) |  |
|  | Khulna | 349 | 69.8 (64.0 to 75.1) |  |
|  | Mymensingh | 135 | 74.2 (63.8 to 82.4) |  |
|  | Rajshahi | 259 | 77.2 (70.9 to 82.5) |  |
|  | Rangpur | 298 | 85.9 (81.1 to 89.7) |  |
|  | Sylhet | 195 | 74.8 (67.9 to 80.6) |  |
| Trimester | First | 806 | 72.6 (69.0 to 76.0) | p=0.038 |
|  | Second | 1,107 | 73.0 (69.9 to 75.9) |  |
|  | Third | 797 | 67.1 (63.1 to 70.9) |  |
| Religion | Muslim or other | 2,503 | 71.2 (69.1 to 73.3) | p=0.986 |
|  | Hindu | 208 | 71.2 (63.0 to 78.1) |  |
| Drinking water source | Piped | 519 | 66.6 (61.8 to 71.0) | p=0.027 |
|  | Tube well | 2,094 | 72.7 (70.4 to 74.9) |  |
|  | Other | 98 | 74.5 (63.2 to 83.3) |  |

### S2 Table. Reproduction of published estimates

| Source and indicator | Published | This analysis |
| --- | --- | --- |
| MICS 2025, Table ATW.15.1S: any anaemia (n=1,940 collected samples) | 53.1 | 53.1 |
| MICS 2025: mild, moderate, severe anaemia | 30.8, 21.5, 0.7 | 30.8, 21.5, 0.7 |
| MICS 2025: any anaemia, first, second, third trimester | 42.0, 52.0, 67.6 | 42.0, 52.0, 67.6 |
| MICS 2025: urban, rural | 56.2, 51.8 | 56.2, 51.8 |
| MICS 2025: Khulna division | 56.3 | 56.5 |
| MICS 2025: other seven divisions | as published | identical to one decimal |
| MICS 2025: mean haemoglobin, g/dL | 10.68 | 10.68 |
| BDHS 2011, STATcompiler: any anaemia in pregnant women, adjusted Hb below 11.0 g/dL (n=352) | 49.6 | 49.6 |

The report counts the six laboratory rejects in the denominator as not anaemic; the main analysis excludes them (53.2%). The report labels severity bands with first and third trimester values but applies the second trimester bands of WHO 2024 to second trimester women, which our figures reproduce.

### S3 Table. Associations with anaemia and haemoglobin, MICS 2025

**A. Crude and minimally adjusted prevalence ratios (Figure 3).** Each characteristic was estimated in its own model, adjusted for the variables listed, chosen in advance as plausible prior causes of that characteristic. Trimester is in every set because the anaemia cut-off depends on it. Complete-case sample, n=1,932. Reference categories as in part B.

| Term | Adjusted for | Any anaemia, crude PR | Any anaemia, minimally adjusted PR | Moderate or severe, crude PR | Moderate or severe, minimally adjusted PR |
| --- | --- | --- | --- | --- | --- |
| Trimester: second | None | 1.24 (1.08 to 1.42) | 1.24 (1.08 to 1.42) | 1.50 (1.11 to 2.01) | 1.50 (1.11 to 2.01) |
| Trimester: third |  | 1.61 (1.41 to 1.84) | 1.61 (1.41 to 1.84) | 2.65 (2.01 to 3.49) | 2.65 (2.01 to 3.49) |
| Age 20-24 | Trimester | 0.87 (0.77 to 0.99) | 0.87 (0.77 to 0.99) | 0.96 (0.73 to 1.27) | 0.96 (0.73 to 1.26) |
| Age 25-29 |  | 0.92 (0.80 to 1.05) | 0.92 (0.80 to 1.05) | 1.12 (0.85 to 1.48) | 1.12 (0.86 to 1.47) |
| Age 30-49 |  | 1.02 (0.90 to 1.17) | 1.01 (0.88 to 1.15) | 1.01 (0.74 to 1.38) | 0.98 (0.72 to 1.32) |
| Previous births: 1 | Trimester, age | 0.98 (0.87 to 1.10) | 0.99 (0.86 to 1.14) | 1.27 (1.00 to 1.62) | 1.24 (0.92 to 1.66) |
| Previous births: 2 or more |  | 1.02 (0.90 to 1.14) | 0.98 (0.82 to 1.17) | 1.09 (0.84 to 1.40) | 1.05 (0.73 to 1.51) |
| Education: secondary | Trimester, age, religion, residence, division | 0.91 (0.81 to 1.03) | 0.92 (0.82 to 1.04) | 0.92 (0.73 to 1.16) | 0.93 (0.74 to 1.16) |
| Education: higher secondary or above |  | 0.92 (0.79 to 1.07) | 0.94 (0.81 to 1.09) | 0.69 (0.50 to 0.94) | 0.67 (0.49 to 0.91) |
| Wealth: second | Trimester, age, education, religion, residence, division | 0.97 (0.84 to 1.12) | 0.99 (0.86 to 1.14) | 0.93 (0.70 to 1.24) | 1.02 (0.77 to 1.35) |
| Wealth: middle |  | 0.84 (0.73 to 0.98) | 0.88 (0.76 to 1.03) | 0.90 (0.67 to 1.20) | 1.10 (0.82 to 1.48) |
| Wealth: fourth |  | 0.97 (0.84 to 1.12) | 1.01 (0.87 to 1.17) | 1.05 (0.79 to 1.41) | 1.30 (0.97 to 1.73) |
| Wealth: richest |  | 1.03 (0.88 to 1.22) | 1.12 (0.94 to 1.33) | 0.91 (0.64 to 1.30) | 1.30 (0.88 to 1.91) |
| Urban residence | Trimester, age, religion, division | 1.08 (0.97 to 1.21) | 1.13 (1.02 to 1.26) | 0.96 (0.77 to 1.20) | 1.04 (0.84 to 1.29) |
| Chattogram | Trimester | 0.89 (0.74 to 1.07) | 0.90 (0.75 to 1.07) | 0.76 (0.53 to 1.10) | 0.77 (0.55 to 1.09) |
| Dhaka |  | 0.97 (0.81 to 1.15) | 1.01 (0.85 to 1.20) | 0.92 (0.65 to 1.31) | 1.01 (0.73 to 1.39) |
| Khulna |  | 1.03 (0.85 to 1.24) | 1.06 (0.88 to 1.27) | 1.00 (0.69 to 1.45) | 1.06 (0.75 to 1.49) |
| Mymensingh |  | 1.00 (0.79 to 1.25) | 1.03 (0.82 to 1.28) | 1.16 (0.76 to 1.77) | 1.22 (0.84 to 1.79) |
| Rajshahi |  | 0.96 (0.78 to 1.18) | 0.99 (0.81 to 1.21) | 0.82 (0.54 to 1.24) | 0.88 (0.59 to 1.31) |
| Rangpur |  | 1.05 (0.88 to 1.26) | 1.06 (0.89 to 1.27) | 1.05 (0.74 to 1.50) | 1.07 (0.76 to 1.49) |
| Sylhet |  | 0.94 (0.76 to 1.17) | 0.95 (0.78 to 1.17) | 0.75 (0.48 to 1.18) | 0.77 (0.49 to 1.20) |
| Hindu religion | Trimester, age, residence, division | 1.29 (1.13 to 1.47) | 1.27 (1.12 to 1.45) | 1.48 (1.10 to 2.00) | 1.47 (1.08 to 1.99) |
| Water: tube well | Trimester, wealth, residence, division | 0.97 (0.86 to 1.11) | 0.99 (0.86 to 1.15) | 0.91 (0.70 to 1.16) | 0.84 (0.64 to 1.12) |
| Water: other |  | 0.91 (0.67 to 1.24) | 0.94 (0.69 to 1.27) | 1.03 (0.61 to 1.75) | 1.11 (0.64 to 1.92) |

**B. Mutually adjusted models.** All models include every term shown (n=1,932). Coefficients are adjusted for one another and should not be read as separate effects. Reference categories: first trimester, age 15-19, no previous birth, primary or no education, poorest quintile, rural, Barishal, Muslim or other religion, piped water. IPW, inverse probability of response weight built from the design weight.

| Term | Any anaemia, PR (blood weight) | Any anaemia, PR (IPW) | Any anaemia, PR (unweighted) | Moderate or severe, PR | Hb difference, g/dL |
| --- | --- | --- | --- | --- | --- |
| Trimester: second | 1.25 (1.10 to 1.43) | 1.19 (1.04 to 1.36) | 1.16 (1.03 to 1.30) | 1.50 (1.13 to 2.01) | -0.81 (-0.97 to -0.65) |
| Trimester: third | 1.63 (1.43 to 1.86) | 1.55 (1.36 to 1.77) | 1.55 (1.39 to 1.73) | 2.69 (2.06 to 3.51) | -0.76 (-0.94 to -0.58) |
| Age 20-24 | 0.86 (0.74 to 1.00) | 0.86 (0.74 to 1.01) | 0.90 (0.79 to 1.02) | 0.93 (0.68 to 1.28) | 0.16 (-0.04 to 0.37) |
| Age 25-29 | 0.89 (0.74 to 1.07) | 0.88 (0.73 to 1.07) | 0.93 (0.80 to 1.10) | 1.10 (0.75 to 1.61) | 0.02 (-0.24 to 0.28) |
| Age 30-49 | 0.95 (0.78 to 1.17) | 0.93 (0.75 to 1.14) | 0.98 (0.82 to 1.16) | 0.92 (0.60 to 1.43) | -0.03 (-0.32 to 0.25) |
| Previous births: 1 | 1.00 (0.87 to 1.16) | 1.03 (0.89 to 1.19) | 1.02 (0.90 to 1.16) | 1.19 (0.87 to 1.61) | -0.02 (-0.22 to 0.18) |
| Previous births: 2 or more | 1.03 (0.85 to 1.24) | 1.04 (0.86 to 1.26) | 1.05 (0.90 to 1.24) | 1.02 (0.70 to 1.50) | -0.00 (-0.26 to 0.25) |
| Education: secondary | 0.92 (0.81 to 1.04) | 0.91 (0.81 to 1.03) | 0.93 (0.84 to 1.04) | 0.90 (0.72 to 1.13) | 0.09 (-0.08 to 0.27) |
| Education: higher secondary or above | 0.93 (0.78 to 1.09) | 0.90 (0.76 to 1.06) | 0.95 (0.82 to 1.09) | 0.62 (0.45 to 0.86) | 0.18 (-0.04 to 0.40) |
| Wealth: second | 0.99 (0.86 to 1.14) | 1.00 (0.87 to 1.16) | 0.98 (0.87 to 1.10) | 1.03 (0.78 to 1.36) | 0.10 (-0.09 to 0.28) |
| Wealth: middle | 0.88 (0.76 to 1.03) | 0.89 (0.77 to 1.04) | 0.92 (0.81 to 1.05) | 1.10 (0.82 to 1.47) | 0.10 (-0.12 to 0.31) |
| Wealth: fourth | 1.01 (0.87 to 1.17) | 1.02 (0.87 to 1.18) | 0.98 (0.86 to 1.12) | 1.27 (0.94 to 1.71) | -0.05 (-0.29 to 0.18) |
| Wealth: richest | 1.12 (0.93 to 1.34) | 1.13 (0.94 to 1.36) | 1.08 (0.92 to 1.27) | 1.24 (0.83 to 1.86) | -0.14 (-0.40 to 0.13) |
| Urban residence | 1.10 (0.98 to 1.23) | 1.11 (0.99 to 1.24) | 1.09 (0.99 to 1.20) | 0.97 (0.78 to 1.22) | -0.08 (-0.24 to 0.07) |
| Chattogram | 0.90 (0.75 to 1.08) | 0.92 (0.77 to 1.10) | 0.95 (0.81 to 1.11) | 0.71 (0.50 to 1.00) | 0.25 (0.00 to 0.50) |
| Dhaka | 0.98 (0.82 to 1.17) | 1.00 (0.83 to 1.19) | 0.98 (0.83 to 1.15) | 0.89 (0.63 to 1.24) | 0.10 (-0.14 to 0.35) |
| Khulna | 1.07 (0.89 to 1.29) | 1.09 (0.91 to 1.31) | 1.13 (0.96 to 1.33) | 0.96 (0.68 to 1.37) | -0.06 (-0.34 to 0.22) |
| Mymensingh | 1.04 (0.83 to 1.30) | 1.08 (0.86 to 1.36) | 0.97 (0.78 to 1.21) | 1.16 (0.80 to 1.69) | -0.16 (-0.57 to 0.25) |
| Rajshahi | 1.01 (0.82 to 1.24) | 1.03 (0.84 to 1.27) | 1.00 (0.82 to 1.21) | 0.84 (0.56 to 1.26) | 0.02 (-0.24 to 0.29) |
| Rangpur | 1.07 (0.89 to 1.29) | 1.08 (0.90 to 1.30) | 1.08 (0.91 to 1.27) | 1.03 (0.74 to 1.43) | -0.16 (-0.41 to 0.09) |
| Sylhet | 0.95 (0.77 to 1.18) | 0.97 (0.78 to 1.21) | 0.96 (0.78 to 1.17) | 0.66 (0.42 to 1.03) | 0.18 (-0.09 to 0.46) |
| Hindu religion | 1.27 (1.11 to 1.44) | 1.26 (1.10 to 1.44) | 1.22 (1.08 to 1.38) | 1.50 (1.12 to 2.01) | -0.30 (-0.52 to -0.09) |
| Water: tube well | 1.00 (0.86 to 1.15) | 1.05 (0.90 to 1.22) | 1.04 (0.91 to 1.18) | 0.91 (0.68 to 1.20) | 0.07 (-0.13 to 0.27) |
| Water: other | 0.92 (0.68 to 1.25) | 0.97 (0.72 to 1.31) | 1.07 (0.85 to 1.34) | 1.13 (0.66 to 1.95) | -0.16 (-0.67 to 0.36) |

### S4 Table. Sensitivity analyses

**A. National prevalence, MICS 2025**

| Analysis | n | Anaemia, % (95% CI) |
| --- | --- | --- |
| Main: blood weight, WHO 2024 cut-offs | 1,933 | 53.2 (50.6 to 55.8) |
| Haemoglobin rounded to 0.1 g/dL | 1,933 | 51.0 (48.4 to 53.6) |
| Excluding weeks 12-15 and 26-29 (near trimester limits) | 1,401 | 52.0 (48.9 to 55.0) |
| Excluding heaped weeks (multiples of 4) | 958 | 53.2 (49.5 to 57.0) |
| Excluding Hb below 6 or above 15 g/dL | 1,921 | 53.3 (50.7 to 55.9) |
| Design weight only (wmweight) | 1,933 | 53.6 (51.0 to 56.1) |
| Inverse probability of response weight | 1,933 | 53.9 (51.2 to 56.5) |
| IPW, trimester 1 | 580 | 44.0 (39.2 to 49.0) |
| IPW, trimester 2 | 811 | 51.8 (47.8 to 55.9) |
| IPW, trimester 3 | 542 | 67.5 (62.4 to 72.1) |

**B. Comparison with 2011 under measurement scenarios.** Adjusted PR for 2025 against 2011 (adjusted as in Table 2). Raising 2025 values mimics a 2025 method that reads lower than the 2011 method. On a grid of offsets in steps of 0.01 g/dL, the 95% CI first included 1 at 0.19 g/dL (PR 1.16, 1.00 to 1.36), and the PR reached 1.0 at 0.38 g/dL (1.00, 0.85 to 1.17).

| Scenario | 2011, % | 2025, % | Adjusted PR (95% CI) | p |
| --- | --- | --- | --- | --- |
| Main (2025 Hb as reported) | 42.9 | 53.2 | 1.33 (1.15 to 1.54) | p<0.001 |
| 2025 Hb rounded to 0.1 g/dL | 42.9 | 51.0 | 1.29 (1.11 to 1.50) | p<0.001 |
| 2025 Hb truncated to 0.1 g/dL | 42.9 | 53.2 | 1.33 (1.15 to 1.54) | p<0.001 |
| 2025 Hb raised by 0.2 g/dL | 42.9 | 44.7 | 1.14 (0.98 to 1.33) | p=0.088 |
| 2025 Hb raised by 0.4 g/dL | 42.9 | 38.4 | 0.98 (0.83 to 1.15) | p=0.790 |

**C. Residence-specific comparison under measurement scenarios.** Stratum-specific models estimated with subpopulation variance; adjusted for trimester, age, previous births, education, division and relative wealth.

| 2025 Hb shift, g/dL | Residence | Adjusted PR 2025 vs 2011 (95% CI) | Interaction p |
| --- | --- | --- | --- |
| 0.0 | Rural | 1.14 (0.97 to 1.35) | p<0.001 |
| 0.0 | Urban | 2.36 (1.62 to 3.42) | p<0.001 |
| 0.2 | Rural | 1.00 (0.84 to 1.18) | p=0.001 |
| 0.2 | Urban | 1.94 (1.31 to 2.87) | p=0.001 |
| 0.4 | Rural | 0.86 (0.72 to 1.03) | p=0.002 |
| 0.4 | Urban | 1.66 (1.12 to 2.46) | p=0.002 |

**D. 2025 against 2011 by subgroup.** Stratum-specific models estimated with subpopulation variance.

| Subgroup | Level | Adjusted PR 2025 vs 2011 (95% CI) | p | Interaction p |
| --- | --- | --- | --- | --- |
| Residence | Rural | 1.14 (0.97 to 1.35) | p=0.108 | p<0.001 |
|  | Urban | 2.36 (1.62 to 3.42) | p<0.001 |  |
| Education | Primary or none | 1.14 (0.93 to 1.39) | p=0.201 | p=0.279 |
|  | Secondary | 1.56 (1.21 to 2.02) | p<0.001 |  |
| Wealth (relative) | Poorer two quintiles | 1.15 (0.95 to 1.39) | p=0.156 | p=0.064 |
|  | Richer three quintiles | 1.50 (1.21 to 1.88) | p<0.001 |  |

Restricting the 2011 sample to women aged 15 years or more did not change any estimate, because the five women aged 14 years had zero weight.

### S5 Table. Composition of the 2011 and 2025 analytic samples

Analytic samples: 352 women in 2011 and 1,933 in 2025. Unweighted n; percentages weighted with each survey weight. | Group | Level | 2011, n | 2011, weighted % | 2025, n | 2025, weighted % | |:-|:-|-:|-:|-:|-:| | Trimester | First | 94 | 25.9 | 580 | 30.7 | | | Second | 140 | 41.3 | 811 | 42.6 | | | Third | 118 | 32.8 | 542 | 26.7 | | Age (years) | 15-19 | 96 | 28.7 | 487 | 25.0 | | | 20-24 | 124 | 35.9 | 596 | 31.0 | | | 25-29 | 87 | 22.0 | 447 | 23.4 | | | 30-49 | 45 | 13.3 | 403 | 20.6 | | Previous births | 0 | 122 | 34.2 | 712 | 37.7 | | | 1 | 106 | 30.6 | 657 | 33.8 | | | 2 or more | 124 | 35.2 | 564 | 28.6 | | Education (years) | 0 to 5 | 172 | 50.2 | 415 | 20.9 | | | 6 to 10 | 160 | 45.7 | 1,093 | 56.5 | | | 11 or more | 20 | 4.1 | 424 | 22.5 | | Residence | Rural | 245 | 77.7 | 1,414 | 70.6 | | | Urban | 107 | 22.3 | 519 | 29.4 | | Division (2011 boundaries) | Barishal | 41 | 5.6 | 207 | 5.7 | | | Chattogram | 60 | 20.6 | 429 | 26.2 | | | Dhaka incl. Mymensingh | 59 | 32.0 | 453 | 31.3 | | | Khulna | 38 | 8.9 | 247 | 9.7 | | | Rajshahi | 36 | 11.1 | 199 | 10.6 | | | Rangpur | 47 | 12.3 | 250 | 9.9 | | | Sylhet | 71 | 9.6 | 148 | 6.7 |

### S6 Table. Mean haemoglobin and anaemia by month of pregnancy

MICS weeks of pregnancy divided by four and rounded; BDHS completed months. Estimates based on fewer than 25 women are unreliable. MICS month 7 includes weeks 26 and 27, which use the second-trimester cut-off.

| Month | Survey | n | Mean Hb, g/dL (95% CI) | Anaemia, % (95% CI) |
| --- | --- | --- | --- | --- |
| 1 | 2011 | 5 | 11.97 (10.90 to 13.04) | 24.5 (3.2 to 76.1) |
| 1 | 2025 | 70 | 11.04 (10.72 to 11.36) | 43.2 (30.2 to 57.2) |
| 2 | 2011 | 47 | 11.66 (11.31 to 12.01) | 30.8 (18.4 to 46.9) |
| 2 | 2025 | 228 | 11.32 (11.13 to 11.52) | 39.5 (32.1 to 47.3) |
| 3 | 2011 | 42 | 11.16 (10.78 to 11.55) | 45.8 (30.8 to 61.5) |
| 3 | 2025 | 282 | 11.16 (10.98 to 11.33) | 44.2 (37.5 to 51.0) |
| 4 | 2011 | 45 | 11.10 (10.76 to 11.45) | 30.7 (17.1 to 48.6) |
| 4 | 2025 | 225 | 10.69 (10.50 to 10.89) | 41.6 (34.7 to 49.0) |
| 5 | 2011 | 41 | 10.97 (10.53 to 11.41) | 37.1 (22.4 to 54.6) |
| 5 | 2025 | 273 | 10.36 (10.22 to 10.51) | 53.5 (46.6 to 60.2) |
| 6 | 2011 | 54 | 10.90 (10.45 to 11.36) | 34.5 (22.0 to 49.6) |
| 6 | 2025 | 246 | 10.25 (10.04 to 10.46) | 58.8 (51.1 to 66.0) |
| 7 | 2011 | 56 | 10.59 (10.21 to 10.97) | 63.5 (48.8 to 76.0) |
| 7 | 2025 | 264 | 10.37 (10.17 to 10.57) | 66.2 (58.3 to 73.3) |
| 8 | 2011 | 52 | 10.76 (10.44 to 11.08) | 52.2 (36.8 to 67.2) |
| 8 | 2025 | 215 | 10.59 (10.39 to 10.79) | 65.7 (58.0 to 72.7) |
| 9 | 2011 | 10 | 10.83 (9.83 to 11.83) | 57.1 (24.3 to 84.7) |
| 9 | 2025 | 130 | 10.45 (10.17 to 10.74) | 66.5 (55.9 to 75.7) |

### S7 Table. Anaemia by division, 2011 and 2025 (2011 boundaries)

Mymensingh division (created in 2015) is merged into Dhaka.

| Division | 2011, n | 2011, % (95% CI) | 2025, n | 2025, % (95% CI) |
| --- | --- | --- | --- | --- |
| Barishal | 41 | 51.1 (31.7 to 70.1) | 207 | 55.3 (47.4 to 63.0) |
| Chattogram | 60 | 39.4 (26.2 to 54.4) | 429 | 49.1 (43.3 to 55.0) |
| Dhaka incl. Mymensingh | 59 | 41.6 (30.4 to 53.8) | 453 | 53.9 (48.9 to 58.7) |
| Khulna | 38 | 37.7 (23.4 to 54.5) | 247 | 56.8 (49.3 to 64.0) |
| Rajshahi | 36 | 49.0 (31.5 to 66.7) | 199 | 53.2 (45.1 to 61.1) |
| Rangpur | 47 | 43.6 (29.7 to 58.5) | 250 | 58.3 (51.3 to 64.9) |
| Sylhet | 71 | 46.5 (33.1 to 60.4) | 148 | 52.1 (43.7 to 60.4) |

### S1 Fig. Participant flow

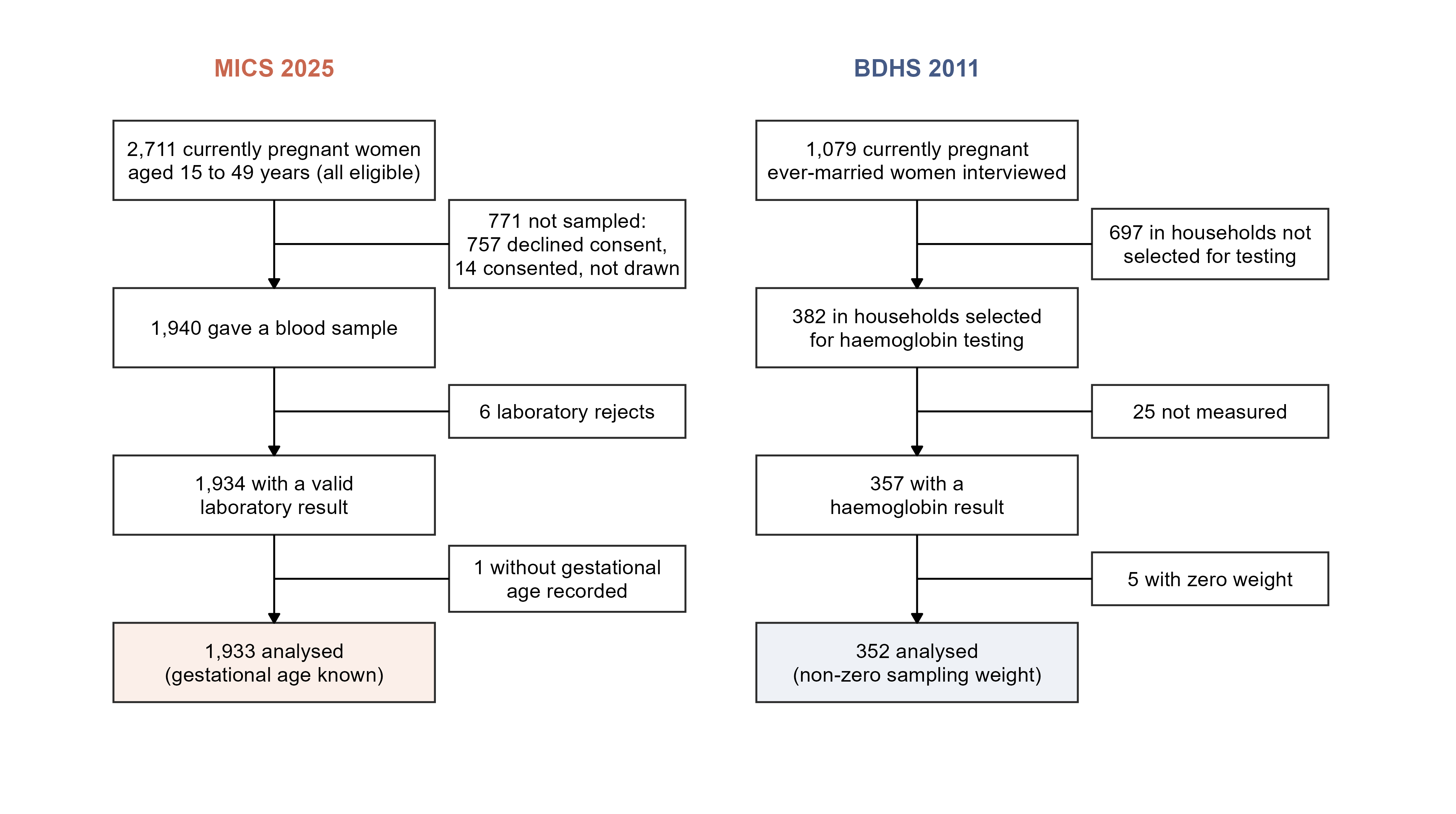

S1 Fig
